# Prenatal Pb exposure is associated with reduced abundance of beneficial gut microbial cliques in late childhood: an investigation using Microbial Co-occurrence Analysis (MiCA)

**DOI:** 10.1101/2023.05.18.23290127

**Authors:** V Midya, JM Lane, C Gennings, LA Torres-Olascoaga, RO Wright, M Arora, MM Téllez-Rojo, S Eggers

## Abstract

**Background:** Many analytical methods used in gut microbiome research focus on either single bacterial taxa or the whole microbiome, ignoring multi-bacteria relationships (microbial cliques). We present a novel analytical approach to identify multiple bacterial taxa within the gut microbiome of children at 9-11 years associated with prenatal Pb exposure.

**Methods:** Data came from a subset of participants (n=123) in the Programming Research in Obesity, Growth, Environment and Social Stressors (PROGRESS) cohort. Pb concentrations were measured in maternal whole blood from the second and third trimesters of pregnancy. Stool samples collected at 9-11 years old underwent metagenomic sequencing to assess the gut microbiome. Using a novel analytical approach, Microbial Co-occurrence Analysis (MiCA), we paired a machine-learning algorithm with randomization-based inference to first identify microbial cliques that were predictive of prenatal Pb exposure and then estimate the association between prenatal Pb exposure and microbial clique abundance.

**Results:** With second-trimester Pb exposure, we identified a 2-taxa microbial clique that included *Bifidobacterium adolescentis* and *Ruminococcus callidus*, and a 3-taxa clique that added *Prevotella clara*. Increasing second-trimester Pb exposure was associated with significantly increased odds of having the 2-taxa microbial clique below the 50^th^ percentile relative abundance (OR=1.03,95%CI[1.01-1.05]). In an analysis of Pb concentration at or above vs. below the United States and Mexico guidelines for child Pb exposure, odds of the 2-taxa clique in low abundance were 3.36(95%CI[1.32-8.51]) and 6.11(95%CI[1.87-19.93]), respectively. Trends were similar with the 3-taxa clique but not statistically significant.

**Discussion:** Using a novel combination of machine-learning and causal-inference, MiCA identified a significant association between second-trimester Pb exposure and reduced abundance of a probiotic microbial clique within the gut microbiome in late childhood. Pb exposure levels at the guidelines for child Pb poisoning in the United States, and Mexico are not sufficient to protect against the potential loss of probiotic benefits.

**Graphical Abstract:** 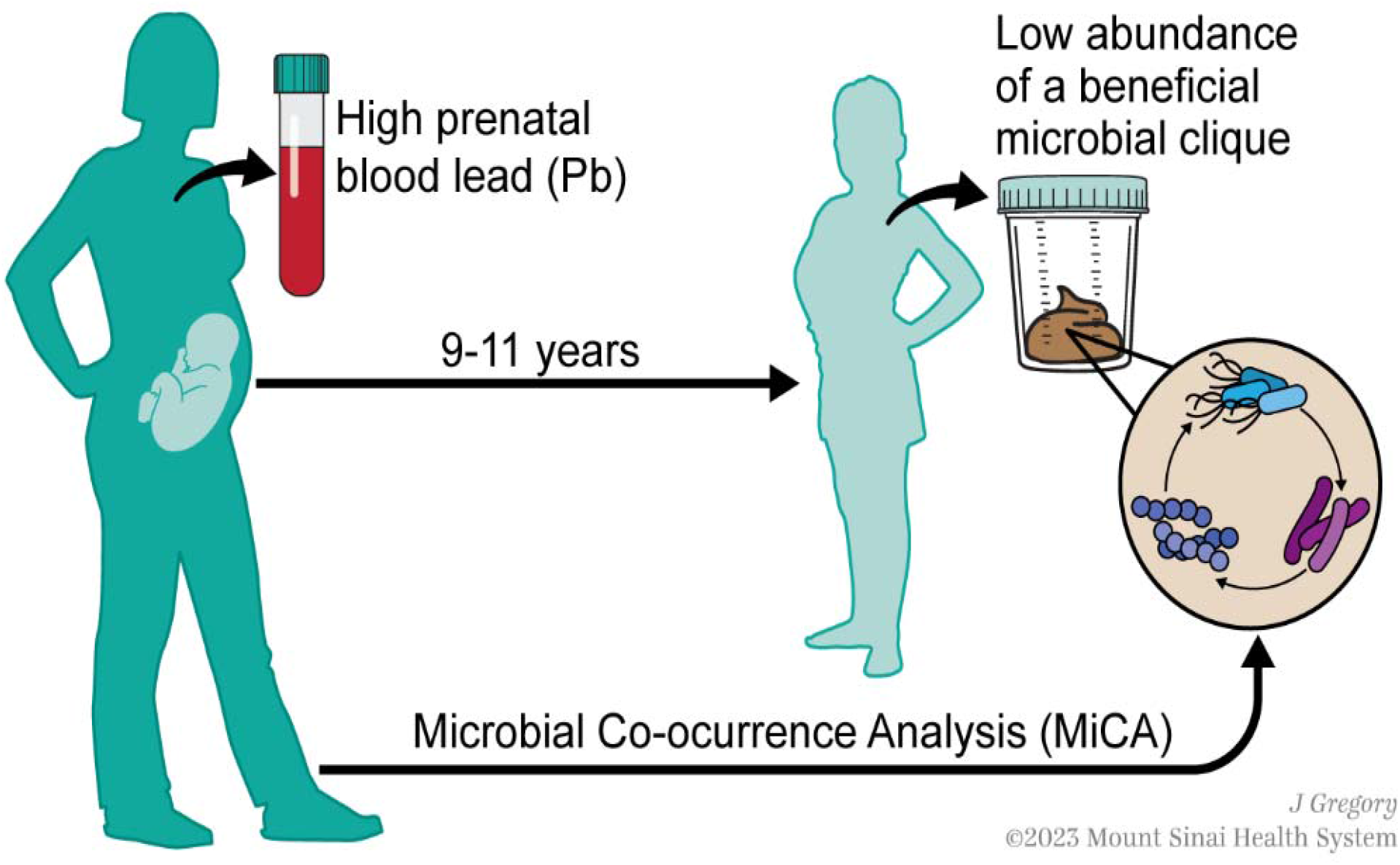

## INTRODUCTION

Human metal exposure has been long recognized as a public health threat, and recent studies suggest that one potential mechanism for their adverse health effects is through the gut microbiome (Claus et al. 2016; Li et al. 2019a). A growing body of animal and human studies have shown that exposure to heavy metals (e.g., As, Cd, and Pb) can alter the gut microbiome composition and metabolic function, reduce diversity, and select for antibiotic resistance (Brabec et al. 2020; Breton et al. 2013; Chi et al. 2017; Eggers et al. 2021; Gao et al. 2017; Li et al. 2019a; Nisanian et al. 2014). Human Pb exposure has been linked to shifts in the microbiome throughout the life course (Eggers et al. 2019; Laue et al. 2020). Sitarik, et al, found that prenatal Pb exposure measured in baby teeth was associated with decreased abundance of several *Bacteroides* species, and an increased abundance of the proinflammatory genus *Collinsella*, in the gut microbiome at 1 and 6 months old (Sitarik et al. 2020). Likewise, Shen, et al, found that prenatal Pb exposure measured in maternal blood was associated with an increased abundance of *Fusobacteriota* in the gut microbiome of children 6-7 years old (Shen et al. 2022). A study out of our group also found that Pb exposure in the second and third trimesters of pregnancy was associated with decreased abundance of several short-chain-fatty-acid-producing bacteria in children 9-11 years of age (Eggers et al. 2023). Taken together, these studies indicate that the prenatal period may be a particularly critical window of Pb exposure on the development of the human gut microbiome in childhood. Moreover, the link between prenatal Pb exposure and increased abundance of pro-inflammatory and potentially pathogenic bacteria, and decreased abundance of probiotic bacteria, suggest that these prenatal Pb exposures may lead to poor health outcomes later in childhood via these alterations to the gut microbiome.

Each of these highlighted studies, and the vast majority of epidemiologic microbiome studies, have used a whole microbiome and/or single taxa approach to investigation. They identified single taxa, or the whole microbiome, as measured using diversity as an indicator associated with a specific exposure or outcome of interest. However, we know from the field of microbial ecology that bacteria (and other microbiome members) biochemically interact with one another at levels between 1-on-1 and the whole microbiome, i.e., groups. For instance, many gut microbes are unable to be cultured in the lab without other bacteria in co-culture (Stewart 2012). In most cases, these bacteria do not need to be co-cultured with everything from the gut microbiome, just one or two others. This group of bacteria within the microbiome, or as we call it, a microbial clique, is missed by the conventional microbiome epidemiology methods. There are studies of the human microbiome that use network analysis to investigate co-occurring microbes within the human microbiome (Dugas et al. 2018; Faust et al. 2012); however, they are based on correlations and can be difficult to use inside of an epidemiologic framework to understand associations and account for confounding variables. Furthermore, the results of network analyses are often difficult to interpret. Thus, the ability to assess associations between exposures or outcomes of epidemiologic interest and microbial cliques within the human microbiome is an important gap in the field. However, finding microbial clique associated with an outcome of interest is challenging because of (1) considerable computational complexity as the number of taxa increases and (2) limitations of sample size in most human microbiome studies. Multiple methods exist where multi-ordered microbial cliques can be pre-specified or hard-coded in the models; however, such strategies might ignore many plausible and informative combinations and could be underpowered due to restrictions on sample size (Gibson 2021; Joubert et al. 2022; Vishal Midya et al. 2023). On the other hand, microbial cliques can be potentially discovered using projection-based dimensionality reduction techniques. However, the interpretations of the final products can be challenging to interpret and are often qualitative (Bellavia). Microbial cliques can be constructed through threshold-based relative abundance, which might aid in interpretation. Such threshold-based construction carries considerable similarity with toxicological threshold-based interactions (Gennings et al. 1997; Hamm et al. 2005; Yeatts et al. 2010). Tree-based machine-learning models can provide a natural and computationally efficient solution to such construction (Vishal Midya et al. 2023). Even with a substantial number of taxa, these models can create multiple threshold-based combinations of taxa predictive of the outcome of interest. Still, the challenge remains in interpretability since most machine-learning models are generally black-box, creating a tension between prediction quality and meaningful interpretability. Moreover, a highly predictive machine-learning model may not be ideal for associations (Shmueli 2010).

In this study, we aim to identify microbial cliques within the gut microbiome of children at 9-11 years old in association with prenatal Pb exposure. To accomplish this, we used a novel statistical approach called Microbial Co-occurrence Analysis (MiCA), which combined interpretable machine learning and causal inference frameworks to first identify microbial cliques, and then test their associations with prenatal Pb exposure.

## METHODS

### Study Design

Data come from the PROGRESS cohort, based out of Mexico City, Mexico. The study enrolled 948 pregnant women who went on to live birth through the Mexican Social Security System. Pregnant women completed study visits in the second trimester (2T), third trimester (3T), and at birth. The offspring were followed and completed study visits every 6 months during infancy and every two years after that. Surveys, physical exams, and psychological and behavioral assessments were conducted at each study visit. Biological specimens, including blood, were also collected. In addition, stool samples were collected for microbiome analysis from a subset of participants (n=123) at ages 9-11 years old. The study protocol for PROGRESS was reviewed and approved by the Institutional Review Board (IRB) at the Icahn School of Medicine at Mount Sinai (ISMMS), and all three committees (Research, Ethics in Research, and Biosafety) included in the IRB at the National Institute of Public Health in Cuernavaca, Mexico.

### Pb Exposure Measurement

Maternal whole blood was drawn during 2T and 3T, and Pb exposure analysis was performed as previously described (Heiss et al. 2020). Pb concentration was measured using inductively coupled plasma mass spectrometry (ICP-MS), at the trace metals laboratory at ISMMS.

### Gut Microbiome Sample Collection and Processing

Microbiome sample collection was conducted as previously described (Eggers et al. 2023). Briefly, whole stool samples were collected at home by participants and stored in the refrigerator until they were picked up by the PROGRESS field team and delivered to ABC Hospital in Mexico City for aliquoting, following the FAST protocol (Romano et al. 2018). Aliquots were stored at -70C, and shipped to the Microbiome Translational Center at ISMMS, where they underwent DNA extraction and library prep in two separate batches (n=50 and n=73). Shotgun metagenomic sequencing was performed for each batch separately using an Illumina HiSeq. Sequencing reads were trimmed for quality using Trimmomatic (Bolger et al. 2014), and bowtie2 (Langmead and Salzberg 2012) was used to remove human reads. MetaPhlAn2 (Truong et al. 2015) and StrainPhlAn (Truong et al. 2017) were then used to determine microbial taxonomy, and HUMAnN2 (Franzosa et al. 2018) was used to profile microbial gene pathways.

### Covariates

Several relevant covariates were considered in this analysis, including child sex, child age at the time of stool sample collection, maternal socio-economic status (SES) during pregnancy, maternal age at birth, maternal body mass index (BMI) during pregnancy, and microbiome analysis batch. Maternal height and weight were measured at 2T and used to calculate BMI. Maternal SES during pregnancy was assessed based on the 1994 Mexican Association of Intelligence Agencies Market and Opinion (AMAI) rule 13*6, where families were categorized into six levels of SES based on 13 questions about household characteristics. Most families in PROGRESS were low to middle SES; therefore, the six levels were condensed into three: lower, middle, and higher (Sanders et al. 2022).

### Statistical Analysis

All analyses were conducted in R version 4.0.3; any two-tailed p-value less than 0.05 was considered statistically significant.

#### Data Processing

Pb concentrations were log_2_ transformed to better meet distributional assumptions. Microbiome count data were converted to relative abundances for all analyses. The analysis included only those taxa with at least 5% relative abundance in both batches to account for analytical batch effects. The relative abundance data were not rescaled to reflect the contribution of the original distribution of the whole taxa. Further, all models were controlled for a batch indicator variable. Further modeling approaches to correct batch effects are described in the following subsection.

#### Microbiome Co-occurrence Analysis (MiCA)

MiCA was conducted in two stages to identify microbial cliques associated with prenatal Pb exposures. The first part of this algorithm used a machine learning-based prediction framework to discover microbial cliques predictive of Pb exposure. The next stage restored the directionality and dived into estimating the association between Pb exposure and the joint-relative abundance of the discovered cliques using a causal inference (or simply classical association-based) framework.

The microbial cliques were searched using repeated hold-out signed-iterated Random Forest (rh-SiRF), where the outcome was prenatal Pb exposure, and the predictors were relative abundances of the selected taxa. The SiRF (Signed Iterative Random Forest) algorithm combined a state-of-the-art predictive tool called “Iterative Random Forests “ with Random Intersection Trees (RIT) to search for combinations of taxa predictive of Pb exposure (Basu et al. 2018; Kumbier et al. 2018; Shah and Meinshausen 2014). Instead of searching for all possible combinations, SiRF can tease out the most prevalent taxa combinations on the decision path. The algorithm begins with a simple random forest (RF) and then sequentially reweights the predictive taxa to fit iterative RFs. From the reweighted RF, decision rules are extracted and fed into a generalization of the RIT to discover microbial cliques from the decision paths. This algorithm introduces a bagging step to assess the “stability “ of the discovered cliques estimated through many bootstrapped iterations. Therefore, the higher the stability of a discovered clique, the better. On top of the SIRF algorithm, we introduced a repeated hold-out step that randomly partitions the data in training and testing sets for better generalizability (Tanner et al. 2019). The whole rh-SiRF algorithm is repeated many times.

The rh-SiRF discovers microbial cliques through decision paths, representing a collective form associated with the outcome rather than a particular functional form. Thus, the discovered cliques include information on directionality and relative abundance thresholds. We fitted the SiRF algorithm in three ways, (1) trained the model on one batch and then tested it on another batch, (2) trained the model on randomly chosen 60% of the data and then tested it on the remaining 40%, and lastly (3) repeated the rh-SiRF algorithm over 300 times with a training and test data partitioning of 60%-40% irrespective of the batches. Microbial cliques were chosen based on having a stability of more than 75%, a prevalence of more than 10%, being common to all data partitioning strategies, and having a higher than random chance of occurrence among the 300 repeated hold-outs (i.e., the relative frequency of occurrence was more than 1%). While fitting the SiRF algorithm on the first two data partitioning strategies (i.e., training on one batch and testing on another and 60%-40% data splitting), we calculated the exposure co-occurrence list to find the important cliques based on mutual co-occurrence. The idea of exposure co-occurrence list follows from the heuristics of distributed word representations (popularly known as word embedding) and is widely applied in tasks related to Natural Language Processing (Li et al. 2015; Lin 2008).

For the next stage of association analysis, the discovered microbial cliques were extracted as indicator functions with respect to their median relative abundances. For example, a microbial clique of A and B, denoted by A-B-, implies that a lower relative abundance of A and B is predictive of prenatal Pb exposure, whereas A+B+ means a higher relative abundance of A and B is predictive of prenatal Pb exposure. For ease of interpretation and generalizability, we converted A-B- to an indicator function with respect to their median relative abundances, i.e., for an individual, the indicator would be non-zero if both the taxa A and B had below median relative abundance; else, it would be zero.

We implemented a randomization-based inference built upon the Rubin causal model to estimate the association between prenatal Pb exposure and the odds of microbial clique abundance below the median. First, a matched-sampling strategy was utilized to obtain similar covariate distributions between the binarized clique – below and above median relative abundance. Given the covariates, we assumed that this approach of covariate-balancing (Greifer 2023) could create potential “exchangeable “ groups so that the clique was hypothetically and randomly assigned to each individual, and those covariates did not confound the clique assignment. Next, due to the small sample size and to prevent discarding a large number of samples, a subclass matching with the propensity score (Stuart et al. 2011) was used to construct similar groups of microbial cliques with above and below-median relative abundance. Finally, love plots of the differences in standardized means in covariates were used to examine the extent of covariate balancing (setting the threshold for the standardized mean difference to 0.1) (Love 2004; Zhang et al. 2019). We used logistic regression with matched microbial clique as the outcome and prenatal Pb concentration as the exposure after adjusting for covariates. Moreover, without relying on asymptotic arguments, we conducted randomization-based inference to construct the null randomization distribution of the test statistic by considering 10^5^ possible exposure assignments and estimated the randomization-based p-value. We also estimated 95% Fisher Confidence Intervals (CIs) based on the randomized p-value under this framework (Imbens and Rubin 2015).

In addition to the previously described steps to eliminate batch effects (5% relative abundance in both batches and multiple SiRF training and testing approaches), we also included the batch indicator in covariate balancing and included batch indicator as a covariate in all models. Any missing data in the covariates or exposures were imputed using the predictive mean matching implementation of the MICE package in R (Buuren and Groothuis-Oudshoorn 2011).

#### Policy relevant Pb Concentration Analysis

We conducted an exploratory analysis using policy-relevant Pb concentration thresholds to estimate the odds of having a below-median relative abundance of the microbial clique at Pb concentrations that were easily interpretable. We dichotomized the sample at the United States guideline level for child Pb poisoning (3.5 ug/dL) (Blood Lead Reference Value | Lead | CDC 2022), the Mexican guideline for child Pb poisoning (5 ug/dL) (DOF - Official Gazette of the Federation) and the study median of prenatal Pb exposure. Odds were estimated for having a below-median relative abundance of each member of the microbial clique with respect to dichotomized Pb exposure (above vs. below the policy-relevant thresholds). Like previous analyses, odds ratios were estimated using logistic regressions after covariate-balancing and subclass-based matching. Since this is an exploratory analysis, we estimated the 95%CIs based on distributional asymptotic arguments. All models were further adjusted by covariates.

#### Gene Pathway Analysis

To interpret the functionality of the cliques, we studied the gene pathways associated (1) with each individual clique member and (2) shared by all clique members, using Venn diagrams. We extracted the gene pathways to understand their joint functionality.

#### Sensitivity Analyses

We conducted multiple sensitivity analyses. First, we repeated the association analyses (1) without the randomization-based causal inference framework (i.e., without any covariate balancing or matching) and (2) without imputing any missing covariate data. Second, the randomization-based causal inference framework was repeated using separate thresholds (25^th^ and 40^th^ percentile) for microbial clique abundance rather than the 50^th^ percentile for each clique member. Third, the models were further adjusted for postnatal child Pb exposure at 12 and 24 months. Fourth, we estimated the Pearson correlations for the taxa within cliques to understand whether relative abundances of the taxa were associated with clique formation. Fifth, similar to the policy-relevant Pb Concentration analysis, we conducted an exploratory study using Pb concentration thresholds to identify the quantile with the highest odds with respect to the indicator of microbial clique abundance. Finally, we dichotomized the sample with gradually increasing percentile thresholds of prenatal Pb exposure, considering all exposures at or above the threshold vs. below the threshold, and sequentially estimated the associations with microbial clique relative abundance all below the median.

## RESULTS

### Study Population

The study population comprised 49 females and 74 males, with an average age of 9.7 years. The 5^th^ and 95^th^ percentiles of observed Pb concentration for 2T and 3T were 10.86 ug/L to 89.18 ug/L and 11.78 ug/L to 77.06 ug/L, respectively. The mean Pb concentration at 2T was 33.6 ug/L and 34.9 ug/L at 3T. Mothers with lower SES during pregnancy were more likely to have higher blood Pb concentrations in both trimesters.

**Table 1.**
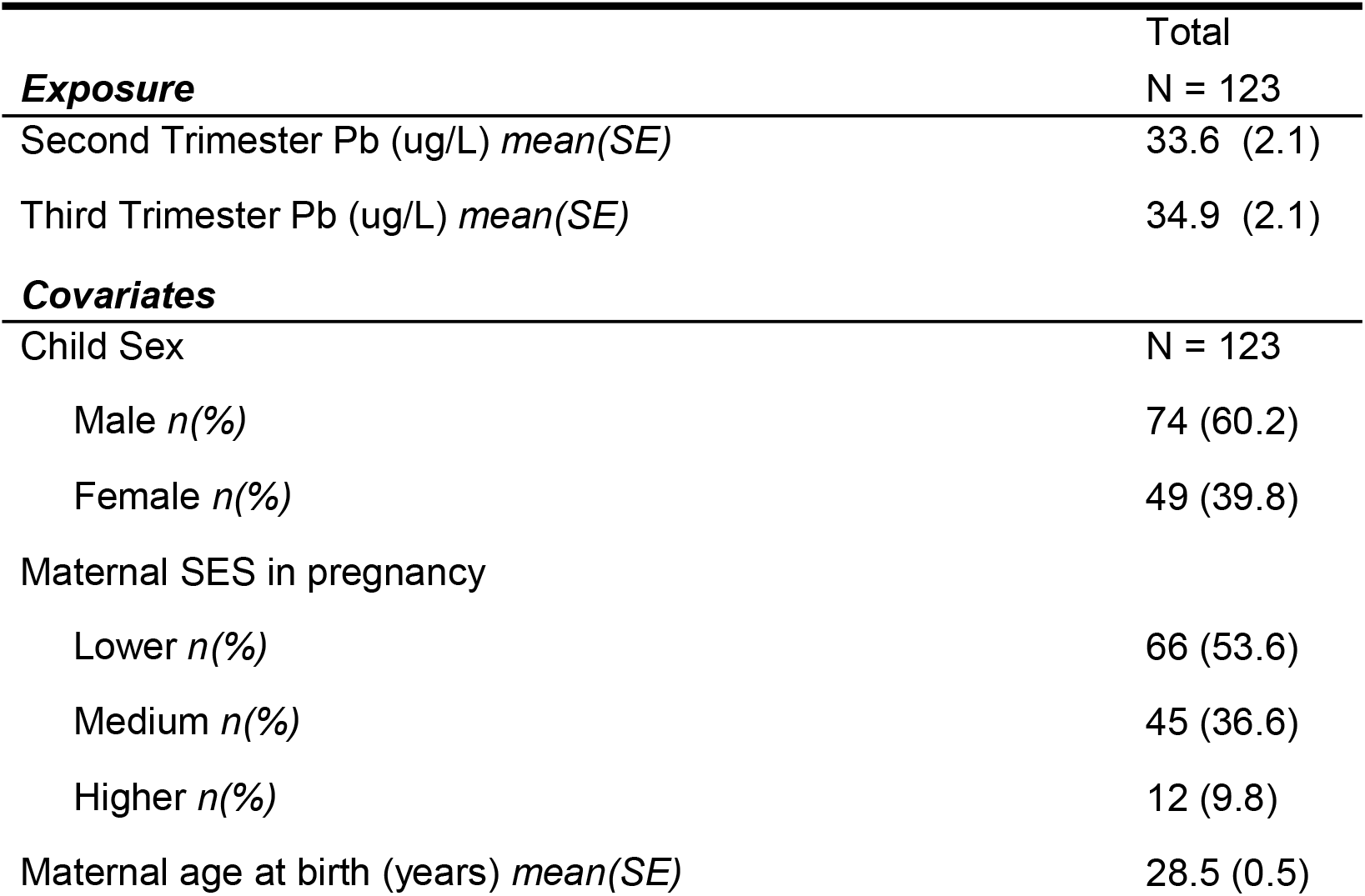
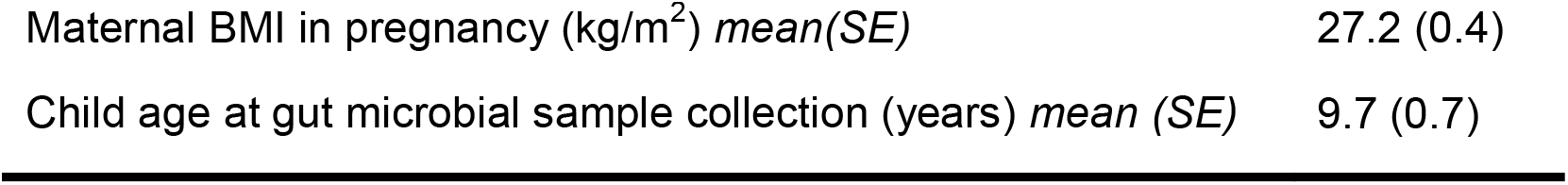
Descriptive statistics of Pb exposure and covariates from the study population.

### MiCA

In the first stage of MiCA, the rh-SiRF identified three separate 2-taxa cliques predictive of Pb concentration in 2T, which included (1) *Bifidobacterium adolescentis* and *Ruminococcus callidus*, (2) *B. adolescentis* and *Paraprevotella clara*, and (3) *R. callidus* and *P. clara*. All three cliques had a stability of more than 75%, a prevalence of more than 10%, and were common to all three data partitioning techniques. Among these three cliques, the clique of *B. adolescentis* and *R. callidus* had the highest score (6/4 and 6/5) in the exposure co-occurrence list, and, therefore, we denoted it as the primary clique (Supplementary Tables 1). Further, based on the commutativity of three cliques, we hypothesized a 3-taxa clique comprising *B. adolescentis, P. clara*, and *R. callidus* (Figure 1). However, for 3T, no clique was common to all three data partitioning (Supplementary Tables 2). In the sections below, we studied the primary 2-taxa clique of *B. adolescentis* and *R. callidus*, and the 3-taxa clique of *B. adolescentis, P. clara*, and *R. callidus*. Lower relative abundance of all taxa in both the cliques was predictive of 2T Pb concentrations, implying that having both or all three of the bacteria co-occur at low or no abundance is predictive of higher 2T Pb concentration. No microbial cliques were identified as predictive of 3T Pb concentration. The detailed tables from the data partitioning for 2T Pb and 3T Pb are presented in Supplementary Tables 1 and 2.

**Figure 1.**
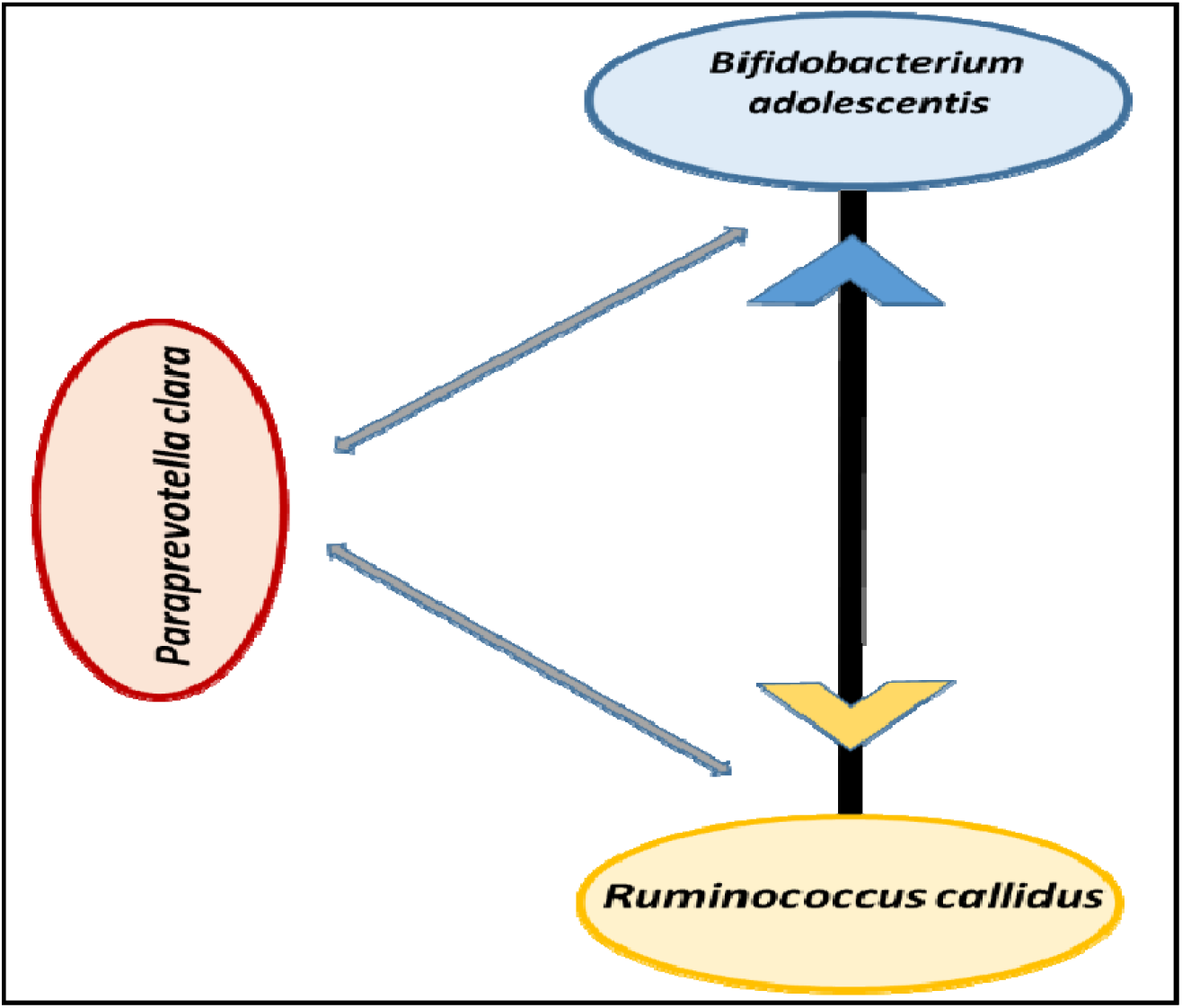
Visual representation of the bacterial taxa in the microbial cliques identified using Microbiome Co-occurrence Analysis (MiCA). The width of the arrows represents the relative frequency of occurrence of the corresponding 2-taxa clique.

In the second stage of MiCA, using a causal inference framework, we found significantly increased odds (OR=1.03, 95% Fisher Confidence Interval (FCIs):[1.01,1.05]), and randomization-based p-value = 0.02] of having both *R. callidus* and *B. adolescentis* below median relative abundance, with increasing Pb concentration in 2T (Figure 2). We also found increased odds (OR=1.02, 95% FCIs:[0.99, 1.04], and randomization-based p-value = 0.16) of all bacteria in the 3-taxa clique (*P. clara, R. callidus* and *B. adolescentis*) having below median relative abundance with increasing 2T Pb concentration, although not statistically significant. Note that covariates were balanced and as well as adjusted in both models. The distribution of propensity scores and the love plot of covariate balancing after subclass matching are presented in Supplementary Figures 1 and 2.

**Figure 2.**
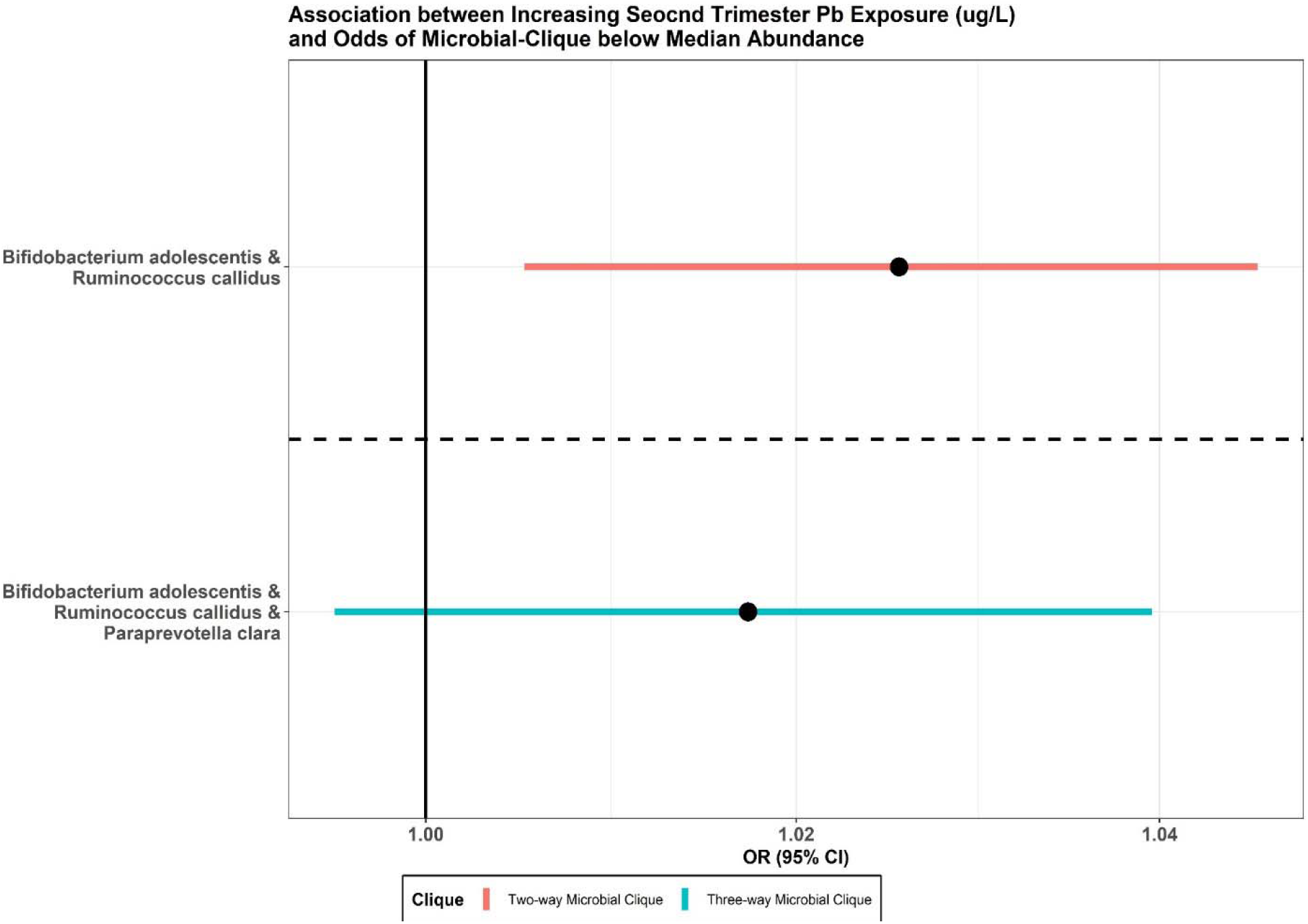
Adjusted odds ratios for below-median abundance (reduced probiotic protection) of all members in 2-taxa (red) and 3-taxa (teal) microbial cliques, with increasing second-trimester prenatal Pb concentration.

### Pb Concentration Thresholds

In an exploratory analysis (Figure 3), we found that those with higher 2T Pb concentration (>= 2.6 ug/dL, i.e., the study median) had higher odds (OR=2.61, 95%CIs:[1.06, 6.45]) of having a below-median relative abundance of the 2-taxa clique (*B. adolescentis* and *R. callidus*). Similarly, those with 2T Pb concentration at or above the United States guideline for child Pb poisoning (>= 3.5 ug/dL) and the Mexican guideline (>= 5 ug/dL) had higher odds (OR=3.36, 95%CIs:[1.32, 8.51] and OR=6.11, 95%CIs:[1.87, 19.93] respectively) of having a below-median relative abundance of the same 2-taxa clique. There is an increasing trend in the odds of below-median relative abundance of the two taxa clique, as the 2T Pb concentration cutoff threshold increases. In other words, children whose mothers had increased 2T Pb concentration, including levels below the US and Mexico guidelines for child Pb poisoning, have increased odds of having a low abundance of the 2-taxa clique late in childhood.

**Figure 3.**
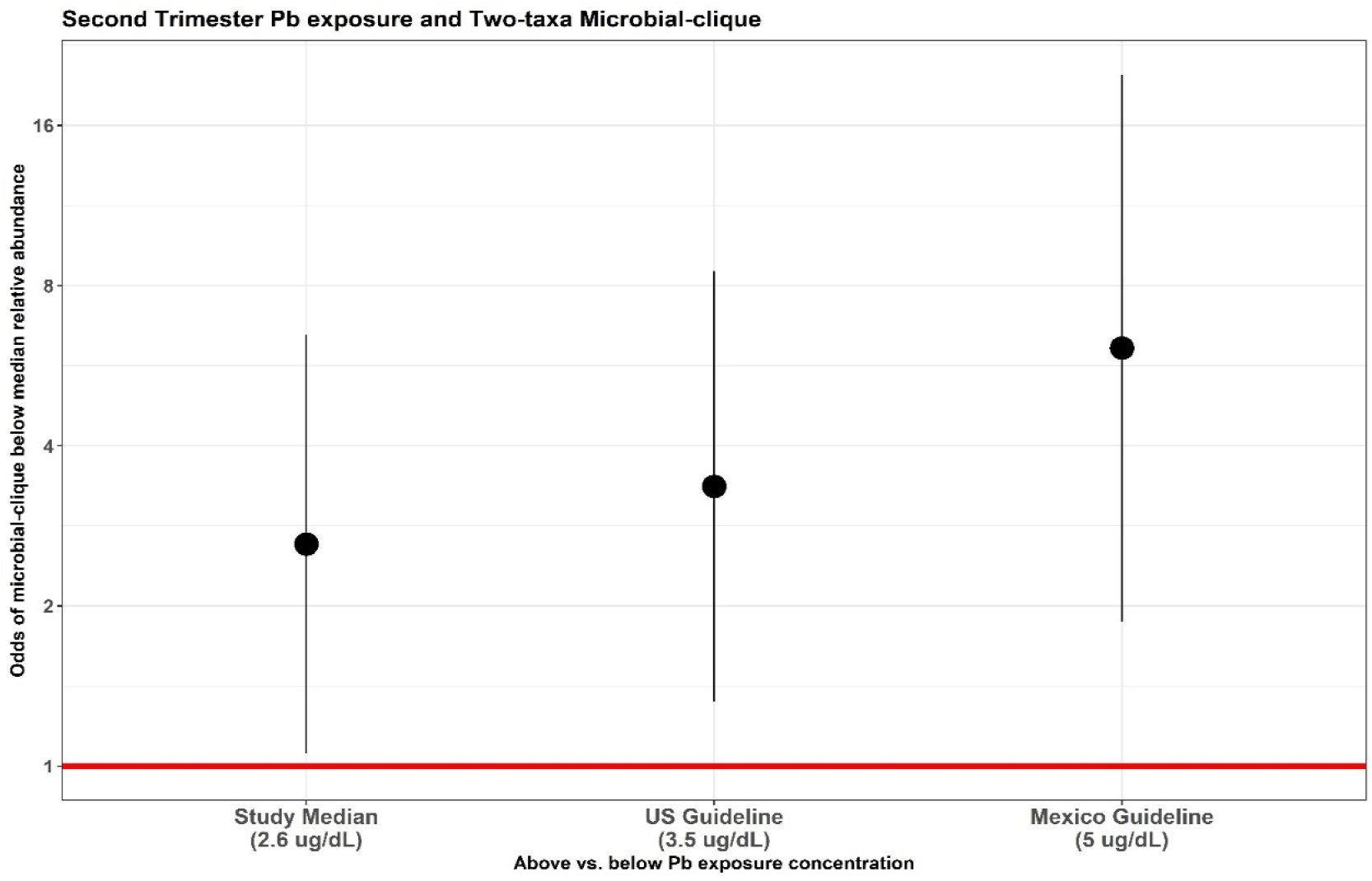
Odds (95% CI) of having below-median relative abundance (reduced probiotic protection) of both members of the 2-taxa clique with respect to 2T Pb concentration above vs. below the cutoff. Pb concentration cutoffs shown are at the study median (2.6 ug/dL), the current United States guideline for child Pb poisoning (3.5 ug/dL), and the current Mexico guideline for child Pb poisoning (5ug/dL). The 5^th^ and 95^th^ percentile of observed Pb concentration was 1.09 ug/dL to 8.92 ug/dL.

### Gene Pathways

In an analysis of the gene pathways belonging to the members of the 2-taxa and 3-taxa microbial cliques identified by MiCA, we examined the gene pathways from each bacterium (Figure 4). The pathways highly abundant in both *B. adolescentis* and *R. callidus* were related to nucleic acid biosynthesis and coenzyme A biosynthesis, essential functions of cellular life. On the other hand, the pathways that were not common between *B. adolescentis* and *R. callidus* in the 2-taxa clique were related more to amino acid biosynthesis and energy metabolism. Similar trends were present when considering *P. clara* as part of the 3-taxa clique. A table of the gene pathways from each taxon can be found in Supplementary Table 3.

**Figure 4.**
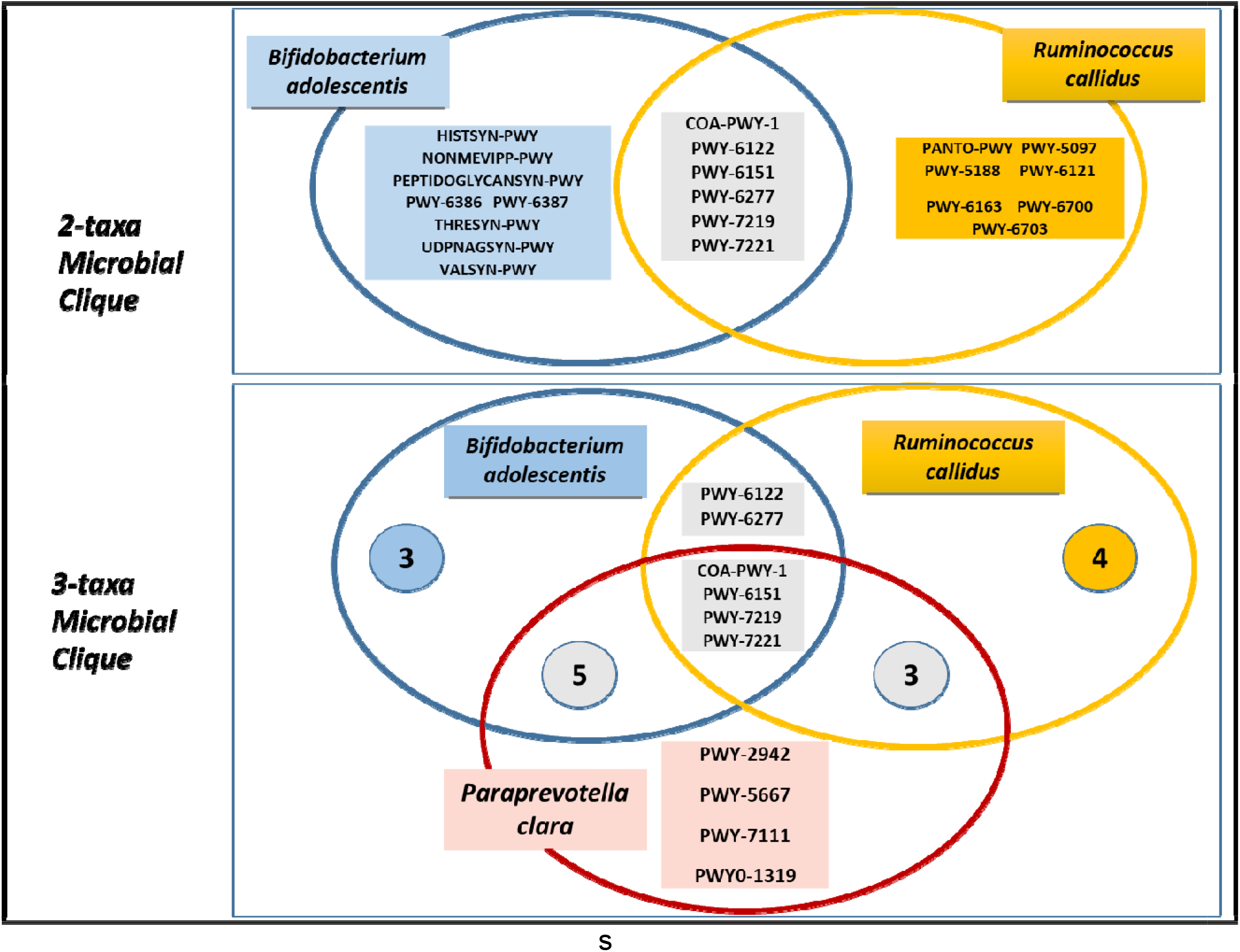
Venn diagrams depicting gene pathways from each taxon within the 2- and 3-taxa microbial cliques. The gene pathways were noted in rectangular boxes. For the 3-taxa microbial clique, numbers in circles denote the number of pathways in that particular subset.

### Exploratory & Sensitivity Analyses

We repeated the association analysis without the causal inference framework and any imputations. The effect sizes did not alter more than 5%, and the model-based asymptotic p-values remained reasonably similar to the randomization-based p-values (Supplementary Table 4). Further, estimates remained practically unchanged (< 5%) after repeating the analysis without imputing any missing covariate (Supplementary Table 5). Under the causal inference framework, the directionality of the associations (ORs) for having both 2-taxa and 3-taxa cliques below 25^th^ or 40^th^ percentile relative abundance remained positively associated with increasing Pb concentration in 2T, although not statistically significant (Supplementary Table 6). After adjusting for child Pb exposure at 12 and 24 months, the associations did not change by more than 5% (Supplementary Table 7). The estimated Pearson ‘s correlation coefficients between the taxa were very small (−0.02 to 0.04), implying relative abundance of the taxa may not be a factor in forming the cliques. Lastly, in an exploratory analysis of 2T Pb concentration, we found that the odds of the 2-taxa clique with below-median relative abundance were highest for those with Pb concentrations in the 50^th^-75^th^ percentile or above (overall aggregated OR=2.76, 95%CI: [1.13, 6.75]). Likewise, the same Pb concentrations were associated with the highest odds of below-median relative abundance for the 3-taxa clique, although the aggregated odds were not statistically significant (mean OR=1.95, 95%CI: [0.73, 5.21]). For two and three taxa cliques, there is an increasing trend in the odds of below-median relative abundance as the cutoff threshold of 2T Pb concentration increases (Supplementary Figure 3).

## DISCUSSION

This study presents a novel approach in microbiome analysis that detects microbial clique(s) predictive of an outcome of interest and estimates the association between that outcome and the clique(s). We used MiCA to identify 2-taxa and 3-taxa microbial cliques in the gut microbiome of children 9-11 years of age, which were negatively causally associated with prenatal Pb exposure. We further explored policy-relevant thresholds for 2T Pb exposure and found significantly increased odds of having below-median relative abundance (reduced probiotic protection) of the 2-taxa microbial clique at and below the current child Pb poisoning guidelines for the United States and Mexico. We also investigated the gene function pathways within the cliques to shed light on their potentially interactive functions.

The microbial clique members, *B. adolescentis, R. callidus*, and *P. clara*, play various beneficial roles within the human gut microbiome. *P. clara* is the most recently identified, with comparatively little known about its health benefits (Morotomi et al. 2009); however, a recent study in dialysis patients found that increased abundance of *P. clara* was associated with reduced constipation (Peng et al. 2023). *R. callidus* is a short-chain-fatty-acid-producing bacteria with anti-inflammatory function (Sánchez-Tapia et al. 2020; Satokari et al. 2014; Sheng et al. 2021). Low abundance of *R. callidus* has been associated with Parkinson ‘s disease (Petrov et al. 2017), colitis and Crohn ‘s disease (Al-Amrah et al. 2023; Kang et al. 2010; Satokari et al. 2014), liver disease, (Sheng et al. 2021) and obesity (Dugas et al. 2018). *B. adolescentis* is a crucial human gut microbe that acts as a starch degrader, Gamma-aminobutyric acid (GABA) producer and helps enhance the intestinal barrier (Altaib et al. 2022; Duranti et al. 2020; Li et al. 2019b; Qian et al. 2022; Wang et al. 2021). *B. adolescentis* is commonly used as a probiotic supplement and has been linked to the prevention and alleviation of many detrimental health conditions, including liver disease (Li et al. 2019b; Long et al. 2021), colitis (Fan et al. 2021; Jang et al. 2019), viral infection (Kim et al. 2014; Li et al. 2016), arthritis (Fan et al. 2020), type 2 diabetes (Qian et al. 2022), anxiety, depression, and other mental health disorders (Jang et al. 2019; Lee et al. 2021). Perhaps its most relevant feature for this analysis, *B. adolescentis* is known to modify the overall composition of the gut microbiome, increasing the abundance of other probiotic or beneficial bacteria within the microbiome, amplifying its beneficial effects (Li et al. 2019b; Qian et al. 2022; Wang et al. 2021). Thus, our finding of *B. adolescentis* as a member of both the 2-taxa and 3-taxa microbial clique, in combination with other potentially beneficial bacterial taxa, is highly consistent with previous literature (Li et al. 2019b; Qian et al. 2022; Wang et al. 2021). When considering the findings of our analysis in the context of this previous evidence, it is clear that prenatal Pb exposure, particularly in the second trimester of pregnancy, has the potential to lead to several detrimental health outcomes, via alterations in the gut microbiome, specifically by reducing the abundance of these 2-taxa and 3-taxa microbial cliques.

The reduced abundance of these probiotic microbial cliques happens not only above the guideline blood Pb concentrations for child Pb poisoning in both Mexico and the United States but even below that at the study median of 2.6ug/dL. Occupational Pb exposure guidelines in the United States only require medical monitoring of employees with blood Pb concentrations above 40ug/dL (Personnel et al. 2012), more than 10x the concentrations observed in this study. Children of mothers with blood Pb levels below the child Pb poisoning guidelines during the second trimester of pregnancy are less likely to have these beneficial gut bacteria in late childhood, and mothers with occupational Pb exposure are likely at even greater risk. Because a reduced abundance of *B. adolescentis* and *R. callidus* have been associated with IBD, liver disease, and reductions in mental health (Al-Amrah et al. 2023; Fan et al. 2021; Lee et al. 2021; Li et al. 2019b; Sheng et al. 2021) the current United States and Mexico guidelines for Pb exposure, while better than past guidelines and procedures, are insufficient to protect against these detrimental health outcomes.

To better understand the potential roles of each taxon within the microbial cliques, we examined the top 20 most abundant gene pathways from each taxon within the cliques. Approximately half of the gene pathways for each taxon were shared with the other clique members, and the other half were unique to that specific taxon. In general, the redundant genes within the clique were key pathways needed for all cellular life, and the unique gene pathways included functions that were more specific to each taxa ‘s metabolism. This indicates that each member of the microbial clique provides unique and potentially complementary functions. Moreover, these taxa are included in the clique not because they are redundant in function and potentially fill the same niche with regard to their association with prenatal Pb exposure but because they are different.

MiCA provides several statistical advantages over other, more traditional microbiome analysis methods (results previously shown using the same data (Eggers et al. 2023)). The amalgamation of interpretable machine-learning algorithm with causal inference tools serves both as predictive as well as associative model. Searching for cliques is difficult when the number of taxa is high (which is the usual scenario), therefore, the usage of machine learning algorithms significantly reduces the computational complexity, and the associative regression models provide interpretability. MiCA does not rely heavily on highly abundant or prevalent taxa within the study samples. As demonstrated in this analysis, MiCA can identify associations with bacteria in low abundance together. MiCA also does not rely on correlations between clique members; thus, cliques can be discovered in association with exposure or outcome of interest even when the taxa within the cliques are not highly correlated within the study sample as a whole. MiCA can also detect cliques in multiple directions with respect to the threshold in a single rh-SiRF analysis. The rh-SiRF step also serves as a major tool for higher prediction accuracy and therefore selects only a few key cliques. Hence, the association tests are highly focused on only a few relationships, reducing the need to correct multiple comparisons. However, the most significant advantage of MiCA is that it analyzes the gut microbiome using a different biological framework than any other epidemiologic analytical tool we know of, i.e., cliques instead of single taxa or the whole microbiome.

While this study presents a novel analytic approach and adds new information about the relationship between Pb exposure and the human gut microbiome, there are some limitations to consider. Limitations of this analysis include using relatively small sample size, with samples processed in multiple batches. However, we took various precautions to reduce batch effects in our estimates and still found statistical significance with small sample size and conservative estimates. Another limitation of this analysis is that we used maternal blood concentration to estimate prenatal Pb exposure in children, which is not a direct exposure estimate. Thus, it is possible that the mechanism of this association may work through maternal exposure rather than prenatal child exposure. For instance, it may be that the maternal Pb exposures alter the maternal gut microbiome during pregnancy, which is then vertically transferred to the offspring at birth, rather than prenatal child Pb exposure priming the child microbiome composition later in childhood. Ultimately, this study is limited in understanding the underlying biochemical mechanism between Pb exposure and the co-occurring bacteria. Further investigation is needed *in vitro* and animal models to better elucidate these mechanisms.

Future analyses could include other biological matrices to estimate Pb exposure, for instance, baby teeth, which can measure direct prenatal exposures starting in the second trimester of pregnancy. We also hope to develop MiCA further to include multiple metal exposures and other potential predictors which may influence the microbiome, including diet and other biological and microbial ecological factors. Future reverse translational studies should be conducted using animal and *in vitro* models to clarify the biochemical mechanisms driving the causal associations identified in this study.

## Data Availability

The data that was used in this study can be made accessible to researchers upon appropriate request with the following restrictions to ensure the privacy of human subjects. Note that access to the data is limited due to a data sharing agreement approved by the IRB at Mount Sinai. Researchers that are interested in accessing PROGRESS data must send their resume/CV as well as CITI training certificates to the IRB chair, Ilene Wilets. They must also submit a data analysis plan to the Principal Investigators for PROGRESS; Robert O. Wright, Martha Tellez-Rojo, and Andrea Baccarelli. Once this process is completed, the PROGRESS data analyst, Nia McRae will send a de-identified dataset via Box, a secure data sharing platform.

## Conflict of Interest

MA is an employee and equity holder of Linus Biotechnology Inc., a start-up company of Mount Sinai Health System. The company develops tools for the detection of autism spectrum disorder and related conditions. The following authors report no competing interests: VM, JML, CG, LATO, ROW, MMTR, SE.

## Funding

This work was supported by the National Institute of Environmental Health Sciences (K99ES032884, P30ES023515, R01ES013744).

## Acknowledgments

The authors would like to acknowledge the entire PROGRESS study team, as well as the participants. We would also like to thank Dr. Jeremiah Faith and the Microbiome Translational Center at the Icahn School of Medicine at Mount Sinai.

## Author Contributions

SE, VM, JML, and CG contributed to conception and design of the study. LATO, MMTR, MA, and ROW contributed to data acquisition. VM performed statistical analysis. SE, VM, CG, and JML contributed to data interpretation. SE wrote the first draft of the manuscript. VM wrote sections of the manuscript. All authors contributed to manuscript revision, read, and approved the submitted version.

## Notes

### Author Declarations

Ethics committee/IRB of the Icahn School of Medicine at Mount Sinai (ISMMS) and all three committees (Research, Ethics in Research, and Biosafety) included in the IRB at the National Institute of Public Health in Cuernavaca, Mexico name gave ethical approval for this work.

