## Supplementary Materials for "Prenatal Pb exposure is associated with reduced abundance of beneficial gut microbial cliques in late childhood: an investigation using Microbial Co-occurrence Analysis (MiCA)"

**Content**

1. Supplementary Table 1: MiCA for Pb exposure at 2T
   1.1 .	Supplementary Table 1a: Train on Batch 2 and Test on Batch 1
   1.2 .	Supplementary Table 1b: Train on 60% and Test on remaining 40%
   1.3 .	Supplementary Table 1c: Repeated Hold-out SiRF (with 60% for training and 40% for testing) repeated over 300 times
2. Supplementary Table 2: MiCA for Pb exposure at 3T
   2.1. Supplementary Table 2a: Train on Batch 2 and Test on Batch 1
   2.2. Supplementary Table 2b: Train on 60% and Test on remaining 40%
   2.3. Supplementary Table 2c: Repeated Hold-out SiRF (with 60% for training and 40% for testing) repeated over 300 times
3. Supplementary Table 3: Gene Pathways
4. Supplementary Table 4: Odds Ratio and 95% CIs from association analysis without any covariate balancing or matching
5. Supplementary Table 5: Association estimates after repeating the analysis without imputing any missing covariate data
6. Supplementary Table 6: Association estimates after cutoff thresholds being set at the 25^th^ or 40^th^ percentile relative abundance
   6.1. Supplementary Table 6a: cutoff thresholds being set at the 40^th^ percentile
   6.2. Supplementary Table 6b: cutoff thresholds being set at the 25^th^ percentile
7. Supplementary Table 7: Association estimates after adjusting for child Pb exposure at 12 and 24 months
8. Supplementary Figure 1: Distribution of propensity scores after covariate balancing
9. Supplementary Figure 2: Love plot of covariate balancing after subclass matching
10. Supplementary Figure 3: Exploratory analysis of 2T Pb concentration

**Supplementary Table 1: MiCA for Pb exposure at 2T**

**Supplementary Table 1a**

**Train on Batch 2 and Test on Batch 1**

| **Cliques** | **Prevalence** | **Precision** | **Stability** | **Exposure co-occurrence frequency** |
| --- | --- | --- | --- | --- |
| **Taxa.2+_Taxa.20-** | 0.113 | 0.617 | 0.988 | 2/4 |
| **Taxa.131-_Taxa.2+** | 0.147 | 0.580 | 1.000 | 6/2 |
| ***Taxa.20-_Taxa.216-*** | **0.112** | **0.533** | **0.956** | **4/2** |
| ***Taxa.131-_Taxa.216-*** | **0.138** | **0.530** | **0.988** | **6/2** |
| ***Taxa.131-_Taxa.20-*** | **0.334** | **0.529** | **1.000** | **6/4** |
| **Taxa.20-_Taxa.43-** | 0.160 | 0.496 | 0.992 | 4/2 |
| **Taxa.131-_Taxa.43-** | 0.193 | 0.501 | 0.992 | 6/2 |

Presenting only those cliques with a stability of more than 0.75 and a prevalence of more than 0.1. See the scientific name of all the Taxa in the Supplementary file “Taxa.names”

**Supplementary Table 1b**

**Train on 60% and Test on remaining 40%**

| **Cliques** | **Prevalence** | **Precision** | **Stability** | **Exposure co-occurrence frequency** |
| --- | --- | --- | --- | --- |
| Taxa.131-_Taxa.22- | 0.102 | 0.520 | 0.880 | 6/2 |
| Taxa.20-_Taxa.22- | 0.111 | 0.516 | 0.964 | 5/2 |
| *Taxa.216-_Taxa.43-* | 0.109 | 0.484 | 0.772 | 3/3 |
| ***Taxa.131-_Taxa.216-*** | **0.206** | **0.485** | **1.000** | **6/3** |
| *Taxa.20-_Taxa.22+* | 0.128 | 0.474 | 0.756 | 5/2 |
| ***Taxa.20-_Taxa.216-*** | ***0.226*** | ***0.482*** | ***1.000*** | ***5/3*** |
| Taxa.131-_Taxa.22+ | 0.142 | 0.471 | 0.792 | 6/2 |
| Taxa.131-_Taxa.43- | 0.202 | 0.469 | 0.988 | 6/3 |
| Taxa.20-_Taxa.43- | 0.221 | 0.466 | 1.000 | 5/3 |
| ***Taxa.131-_Taxa.20-*** | ***0.342*** | ***0.476*** | ***1.000*** | ***6/5*** |

Presenting only those cliques with a stability of more than 0.75 and a prevalence of more than 0.1. See the scientific name of all the Taxa in the Supplementary file “Taxa.names”

**Supplementary Table 1c**

**Repeated Hold-out SiRF (with 60% for training and 40% for testing) repeated over 300 times**

| **Cliques** | **Frequency of occurrence (in %)** |
| --- | --- |
| Taxa.131-_Taxa.5- | 7.9 |
| Taxa.5-_Taxa.94- | 4.9 |
| Taxa.5-_Taxa.61- | 3.9 |
| Taxa.131-_Taxa.42- | 3.6 |
| Taxa.131-_Taxa.61- | 3.6 |
| Taxa.131-_Taxa.94- | 3.6 |
| Taxa.1-_Taxa.131- | 3 |
| Taxa.1-_Taxa.5- | 3 |
| Taxa.30-_Taxa.5- | 3 |
| Taxa.131-_Taxa.2- | 2.6 |
| ***Taxa.131-_Taxa.216-*** | ***2.6*** |
| ***Taxa.131-_Taxa.20-*** | ***2.3*** |
| Taxa.131-_Taxa.76- | 2.3 |
| Taxa.2-_Taxa.5- | 2.3 |
| Taxa.42-_Taxa.5- | 2.3 |
| Taxa.131-_Taxa.30- | 2 |
| Taxa.34-_Taxa.5- | 2 |
| Taxa.42-_Taxa.94- | 2 |
| Taxa.5-_Taxa.76- | 2 |
| Taxa.1-_Taxa.216- | 1.6 |
| Taxa.20-_Taxa.5- | 1.6 |
| Taxa.21-_Taxa.5- | 1.6 |
| Taxa.34-_Taxa.94- | 1.6 |
| Taxa.76-_Taxa.94- | 1.6 |
| Taxa.1-_Taxa.20- | 1.3 |
| Taxa.131-_Taxa.21- | 1.3 |
| Taxa.21-_Taxa.94- | 1.3 |
| Taxa.61-_Taxa.94- | 1.3 |
| Taxa.1-_Taxa.30- | 1 |
| Taxa.1-_Taxa.34- | 1 |
| Taxa.2-_Taxa.61- | 1 |
| ***Taxa.20-_Taxa.216-*** | ***1*** |
| Taxa.21-_Taxa.42- | 1 |
| Taxa.216-_Taxa.5- | 1 |
| Taxa.30-_Taxa.94- | 1 |
| Taxa.42-_Taxa.76- | 1 |
| Taxa.1-_Taxa.42- | 0.7 |
| Taxa.1-_Taxa.61- | 0.7 |
| Taxa.1-_Taxa.76- | 0.7 |
| Taxa.131-_Taxa.2+ | 0.7 |
| Taxa.131-_Taxa.52- | 0.7 |
| Taxa.2-_Taxa.20- | 0.7 |
| Taxa.2-_Taxa.216- | 0.7 |
| Taxa.20-_Taxa.61- | 0.7 |
| Taxa.21-_Taxa.30- | 0.7 |
| Taxa.21-_Taxa.61- | 0.7 |
| Taxa.216-_Taxa.42- | 0.7 |
| Taxa.34-_Taxa.42- | 0.7 |
| Taxa.5-_Taxa.52- | 0.7 |
| Taxa.1-_Taxa.22- | 0.3 |
| Taxa.1-_Taxa.52- | 0.3 |
| Taxa.1-_Taxa.94- | 0.3 |
| Taxa.120-_Taxa.61- | 0.3 |
| Taxa.120-_Taxa.94- | 0.3 |
| Taxa.131-_Taxa.16- | 0.3 |
| Taxa.131-_Taxa.22- | 0.3 |
| Taxa.131-_Taxa.26- | 0.3 |
| Taxa.131-_Taxa.28- | 0.3 |
| Taxa.131-_Taxa.3- | 0.3 |
| Taxa.131-_Taxa.7- | 0.3 |
| Taxa.131-_Taxa.8+ | 0.3 |
| Taxa.16-_Taxa.42- | 0.3 |
| Taxa.16-_Taxa.5- | 0.3 |
| Taxa.16-_Taxa.61- | 0.3 |
| Taxa.2-_Taxa.42- | 0.3 |
| Taxa.2-_Taxa.52- | 0.3 |
| Taxa.2+_Taxa.42- | 0.3 |
| Taxa.2+_Taxa.61- | 0.3 |
| Taxa.20-_Taxa.34- | 0.3 |
| Taxa.21-_Taxa.76- | 0.3 |
| Taxa.216-_Taxa.30- | 0.3 |
| Taxa.216-_Taxa.61- | 0.3 |
| Taxa.26-_Taxa.61- | 0.3 |
| Taxa.28-_Taxa.5- | 0.3 |
| Taxa.3-_Taxa.5- | 0.3 |
| Taxa.30-_Taxa.34- | 0.3 |
| Taxa.30-_Taxa.61- | 0.3 |
| Taxa.42-_Taxa.43- | 0.3 |
| Taxa.42-_Taxa.61- | 0.3 |
| Taxa.43-_Taxa.5- | 0.3 |
| Taxa.5-_Taxa.7- | 0.3 |
| Taxa.5-_Taxa.8+ | 0.3 |
| Taxa.52-_Taxa.94- | 0.3 |
| Taxa.61-_Taxa.8+ | 0.3 |

See the scientific name of all the Taxa in the Supplementary file “Taxa.names”

**Supplementary Table 2: MiCA for Pb exposure at 3T**

**Supplementary Table 2a**

**Train on Batch 2 and Test on Batch 1**

No clique was found with stability of more than 0.75 and a prevalence of more than 0.1

**Supplementary Table 2b**

**Train on 60% and Test on remaining 40%**

| **Cliques** | **Prevalence** | **Precision** | **Stability** | **Exposure co-occurrence frequency** |
| --- | --- | --- | --- | --- |
| Taxa.21-_Taxa.5- | 0.191 | 0.575 | 1.000 | 3/3 |
| Taxa.21-_Taxa.22- | 0.125 | 0.561 | 0.988 | 3/3 |
| Taxa.5-_Taxa.61- | 0.195 | 0.523 | 1.000 | 3/3 |
| Taxa.21-_Taxa.61- | 0.171 | 0.525 | 1.000 | 3/3 |
| Taxa.22-_Taxa.5- | 0.179 | 0.502 | 1.000 | 3/3 |
| Taxa.22-_Taxa.61- | 0.154 | 0.479 | 0.996 | 3/3 |

Presenting only those cliques with a stability of more than 0.75 and a prevalence of more than 0.1. See the scientific name of all the Taxa in the Supplementary file “Taxa.names”

**Supplementary Table 2c**

**Repeated Hold-out SiRF (with 60% for training and 40% for testing) repeated over 300 times**

| **Cliques** | **Frequency of occurrence (in %)** |
| --- | --- |
| Taxa.22-_Taxa.61- | 13 |
| Taxa.21-_Taxa.61- | 8 |
| Taxa.21-_Taxa.22- | 7 |
| Taxa.5-_Taxa.61- | 6 |
| Taxa.111-_Taxa.61- | 5 |
| Taxa.111-_Taxa.22- | 4 |
| Taxa.21-_Taxa.5- | 4 |
| Taxa.22-_Taxa.5- | 4 |
| Taxa.2+_Taxa.61- | 2 |
| Taxa.20-_Taxa.22- | 2 |
| Taxa.20-_Taxa.61- | 2 |
| Taxa.22-_Taxa.31- | 2 |
| Taxa.111-_Taxa.21- | 2 |
| Taxa.2+_Taxa.22- | 2 |
| Taxa.22-_Taxa.30- | 2 |
| Taxa.42-_Taxa.61- | 2 |
| Taxa.111-_Taxa.5- | 1 |
| Taxa.20-_Taxa.30- | 1 |
| Taxa.30-_Taxa.5- | 1 |
| Taxa.30-_Taxa.61- | 1 |
| Taxa.31-_Taxa.61- | 1 |
| Taxa.61-_Taxa.94- | 1 |
| Taxa.111-_Taxa.42- | 1 |
| Taxa.111-_Taxa.49- | 1 |
| Taxa.111-_Taxa.6- | 1 |
| Taxa.131-_Taxa.20- | 1 |
| Taxa.131-_Taxa.22- | 1 |
| Taxa.131-_Taxa.30- | 1 |
| Taxa.131-_Taxa.5- | 1 |
| Taxa.131-_Taxa.9+ | 1 |
| Taxa.131-_Taxa.94- | 1 |
| Taxa.16-_Taxa.31- | 1 |
| Taxa.16-_Taxa.61- | 1 |
| Taxa.2+_Taxa.20- | 1 |
| Taxa.2+_Taxa.5- | 1 |
| Taxa.2+_Taxa.94- | 1 |
| Taxa.20-_Taxa.21- | 1 |
| Taxa.20-_Taxa.43- | 1 |
| Taxa.20-_Taxa.8+ | 1 |
| Taxa.20-_Taxa.94- | 1 |
| Taxa.21-_Taxa.31- | 1 |
| Taxa.21-_Taxa.48- | 1 |
| Taxa.21-_Taxa.49- | 1 |
| Taxa.21-_Taxa.6- | 1 |
| Taxa.21+_Taxa.22- | 1 |
| Taxa.21+_Taxa.31- | 1 |
| Taxa.22-_Taxa.42- | 1 |
| Taxa.22-_Taxa.43- | 1 |
| Taxa.22-_Taxa.48- | 1 |
| Taxa.22-_Taxa.7- | 1 |
| Taxa.22-_Taxa.9+ | 1 |
| Taxa.22+_Taxa.31- | 1 |
| Taxa.22+_Taxa.61- | 1 |
| Taxa.3-_Taxa.42- | 1 |
| Taxa.3-_Taxa.5- | 1 |
| Taxa.3-_Taxa.61- | 1 |
| Taxa.3-_Taxa.94- | 1 |
| Taxa.30-_Taxa.94- | 1 |
| Taxa.31-_Taxa.5- | 1 |
| Taxa.42-_Taxa.5- | 1 |
| Taxa.42-_Taxa.94- | 1 |
| Taxa.49-_Taxa.5- | 1 |
| Taxa.49-_Taxa.61- | 1 |
| Taxa.5-_Taxa.6- | 1 |
| Taxa.61-_Taxa.7- | 1 |
| Taxa.61-_Taxa.8+ | 1 |

See the scientific name of all the Taxa in the Supplementary file “Taxa.names”

**Supplementary Table 3: Gene Pathways**

| **Taxon** | **Gene Pathway** |
| --- | --- |
| **Bifidobacterium adolescentis** | COA-PWY-1: coenzyme A biosynthesis II (mammalian); HISTSYN-PWY: L-histidine biosynthesis; NONMEVIPP-PWY: methylerythritol phosphate pathway I; PEPTIDOGLYCANSYN-PWY: peptidoglycan biosynthesis I (meso-diaminopimelate containing); PWY-6122: 5-aminoimidazole ribonucleotide biosynthesis II; PWY-6151: S-adenosyl-L-methionine cycle I; PWY-6277: superpathway of 5-aminoimidazole ribonucleotide biosynthesis; PWY-6386: UDP-N-acetylmuramoyl-pentapeptide biosynthesis II (lysine-containing); PWY-6387: UDP-N-acetylmuramoyl-pentapeptide biosynthesis I (meso-diaminopimelate containing); PWY-7219: adenosine ribonucleotides de novo biosynthesis; PWY-7221: guanosine ribonucleotides de novo biosynthesis; THRESYN-PWY: superpathway of L-threonine biosynthesis; UDPNAGSYN-PWY: UDP-N-acetyl-D-glucosamine biosynthesis I; VALSYN-PWY: L-valine biosynthesis |
| **Paraprevotella clara** | COA-PWY-1: coenzyme A biosynthesis II (mammalian); NONMEVIPP-PWY: methylerythritol phosphate pathway I; PANTO-PWY: phosphopantothenate biosynthesis I; PEPTIDOGLYCANSYN-PWY: peptidoglycan biosynthesis I (meso-diaminopimelate containing); PWY-2942: L-lysine biosynthesis III; PWY-5097: L-lysine biosynthesis VI; PWY-5667: CDP-diacylglycerol biosynthesis I; PWY-6151: S-adenosyl-L-methionine cycle I; PWY-6386: UDP-N-acetylmuramoyl-pentapeptide biosynthesis II (lysine-containing); PWY-6387: UDP-N-acetylmuramoyl-pentapeptide biosynthesis I (meso-diaminopimelate containing); PWY-6700: queuosine biosynthesis; PWY-7111: pyruvate fermentation to isobutanol (engineered); PWY-7219: adenosine ribonucleotides de novo biosynthesis; PWY-7221: guanosine ribonucleotides de novo biosynthesis; PWY0-1319: CDP-diacylglycerol biosynthesis II; VALSYN-PWY: L-valine biosynthesis |
| **Ruminococcus callidus** | COA-PWY-1: coenzyme A biosynthesis II (mammalian); PANTO-PWY: phosphopantothenate biosynthesis I; PWY-5097: L-lysine biosynthesis VI; PWY-5188: tetrapyrrole biosynthesis I (from glutamate); PWY-6121: 5-aminoimidazole ribonucleotide biosynthesis I; PWY-6122: 5-aminoimidazole ribonucleotide biosynthesis II; PWY-6151: S-adenosyl-L-methionine cycle I; PWY-6163: chorismate biosynthesis from 3-dehydroquinate; PWY-6277: superpathway of 5-aminoimidazole ribonucleotide biosynthesis; PWY-6700: queuosine biosynthesis; PWY-6703: preQ0 biosynthesis; PWY-7219: adenosine ribonucleotides de novo biosynthesis; PWY-7221: guanosine ribonucleotides de novo biosynthesis |

**Supplementary Table 4: Odds Ratio and 95% CIs from association analysis without any covariate balancing or matching**

| **Outcome** | **Odds Ratio** | **95% CI** | **p-value** |
| --- | --- | --- | --- |
| Below-median relative abundance of the 2-taxa microbial clique | 1.02 | (1.00, 1.04) | 0.03 |
| Below-median relative abundance of the 3-taxa microbial clique | 1.01 | (0.99, 1.03) | 0.17 |

**Supplementary Table 5: Association estimates after repeating the analysis without imputing any missing covariate data**

| **Outcome** | **Odds Ratio** | **95% CI** | **p-value** |
| --- | --- | --- | --- |
| Below-median relative abundance of the 2-taxa microbial clique | 1.02 | (1.00, 1.04) | 0.04 |
| Below-median relative abundance of the 3-taxa microbial clique | 1.01 | (0.99, 1.03) | 0.28 |

**Supplementary Table 6: Association estimates after cutoff thresholds being set at the 25^th^ or 40^th^ percentile relative abundance**

**Supplementary Table 6a: Cutoff thresholds being set at the 40^th^ percentile**

| **Outcome** | **Odds Ratio** | **95% CI** | **p-value** |
| --- | --- | --- | --- |
| Below-median relative abundance of the 2-taxa microbial clique | 1.02 | (1.00, 1.04) | 0.11 |
| Below-median relative abundance of the 3-taxa microbial clique | 1.01 | (0.99, 1.03) | 0.34 |

**Supplementary Table 6b: Cutoff thresholds being set at the 25^th^ percentile**

| **Outcome** | **Odds Ratio** | **95% CI** | **p-value** |
| --- | --- | --- | --- |
| Below-median relative abundance of the 2-taxa microbial clique | 1.01 | (0.99, 1.04) | 0.26 |
| Below-median relative abundance of the 3-taxa microbial clique | 1.00 | (0.98, 1.04) | 0.63 |

**Supplementary Table 7: Association estimates after adjusting for child Pb exposure at 12 and 24 months**

| **Outcome** | **Odds Ratio** | **95% CI** | **p-value** |
| --- | --- | --- | --- |
| Below-median relative abundance of the 2-taxa microbial clique | 1.02 | (1.00, 1.04) | 0.04 |
| Below-median relative abundance of the 3-taxa microbial clique | 1.01 | (0.99, 1.04) | 0.18 |

**Supplementary Figure 1: Distribution of propensity scores after covariate balancing**

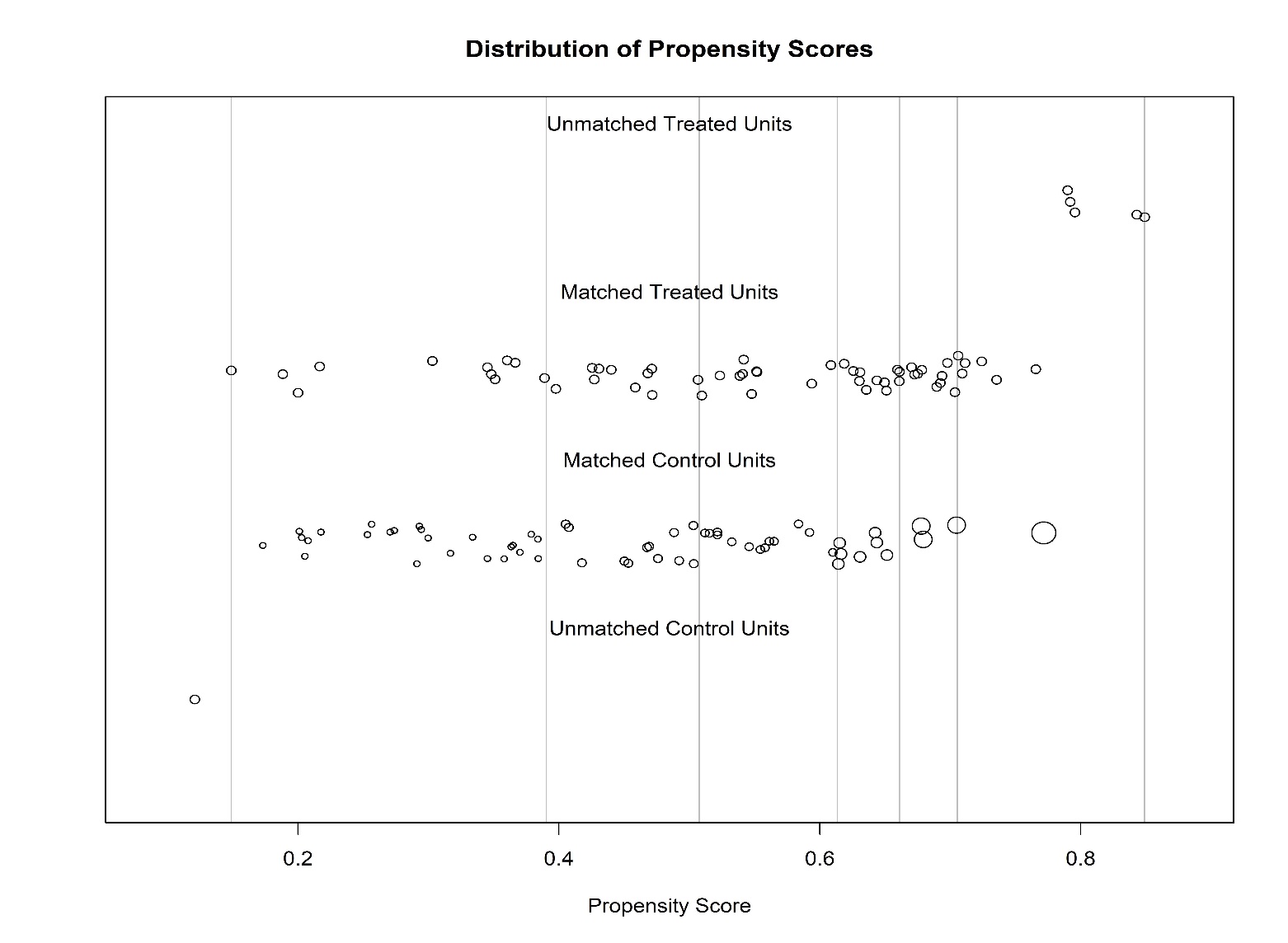

**Supplementary Figure 2: Love plot of covariate balancing after subclass matching**

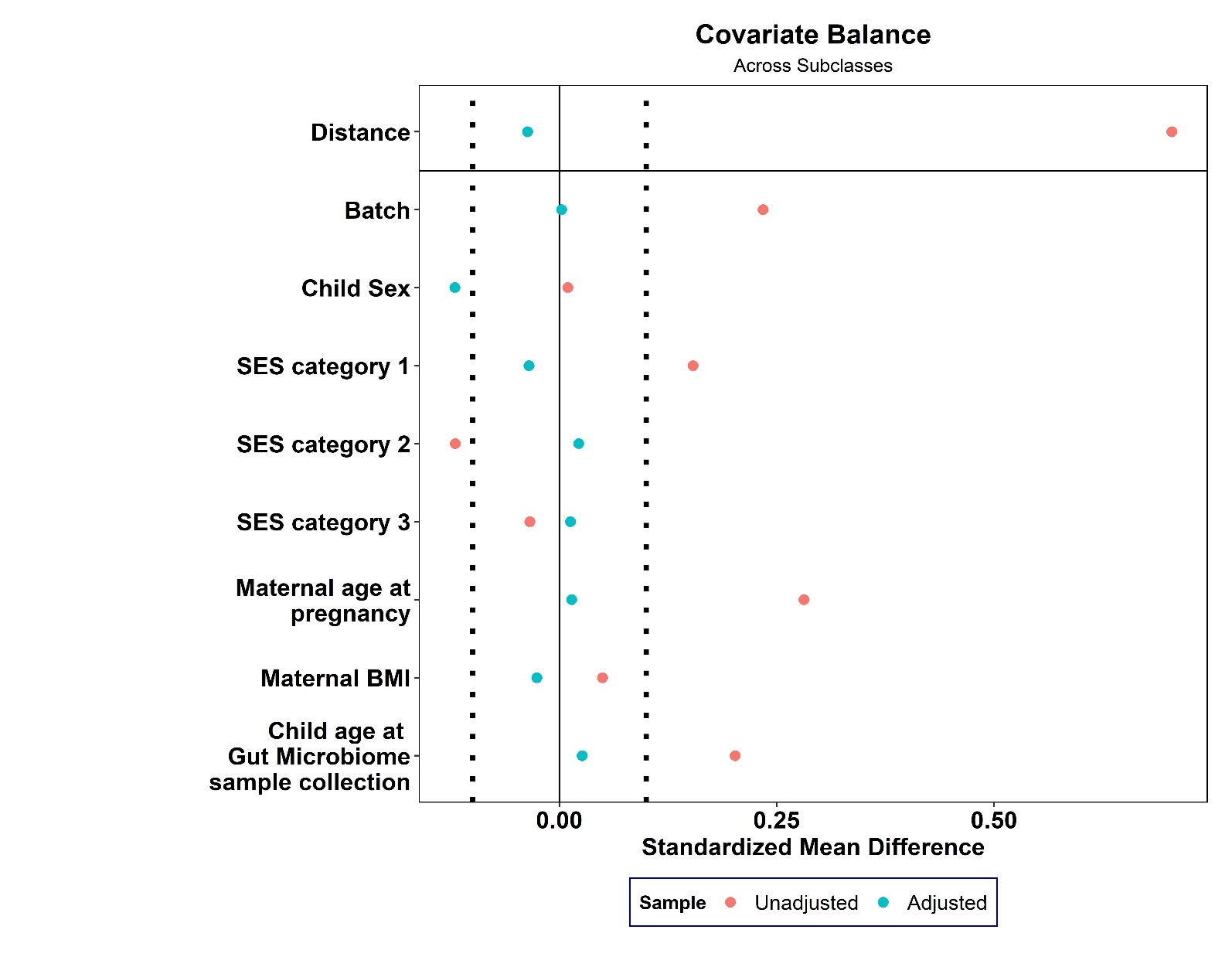

**Supplementary Figure 3: Exploratory analysis of 2T Pb concentration**

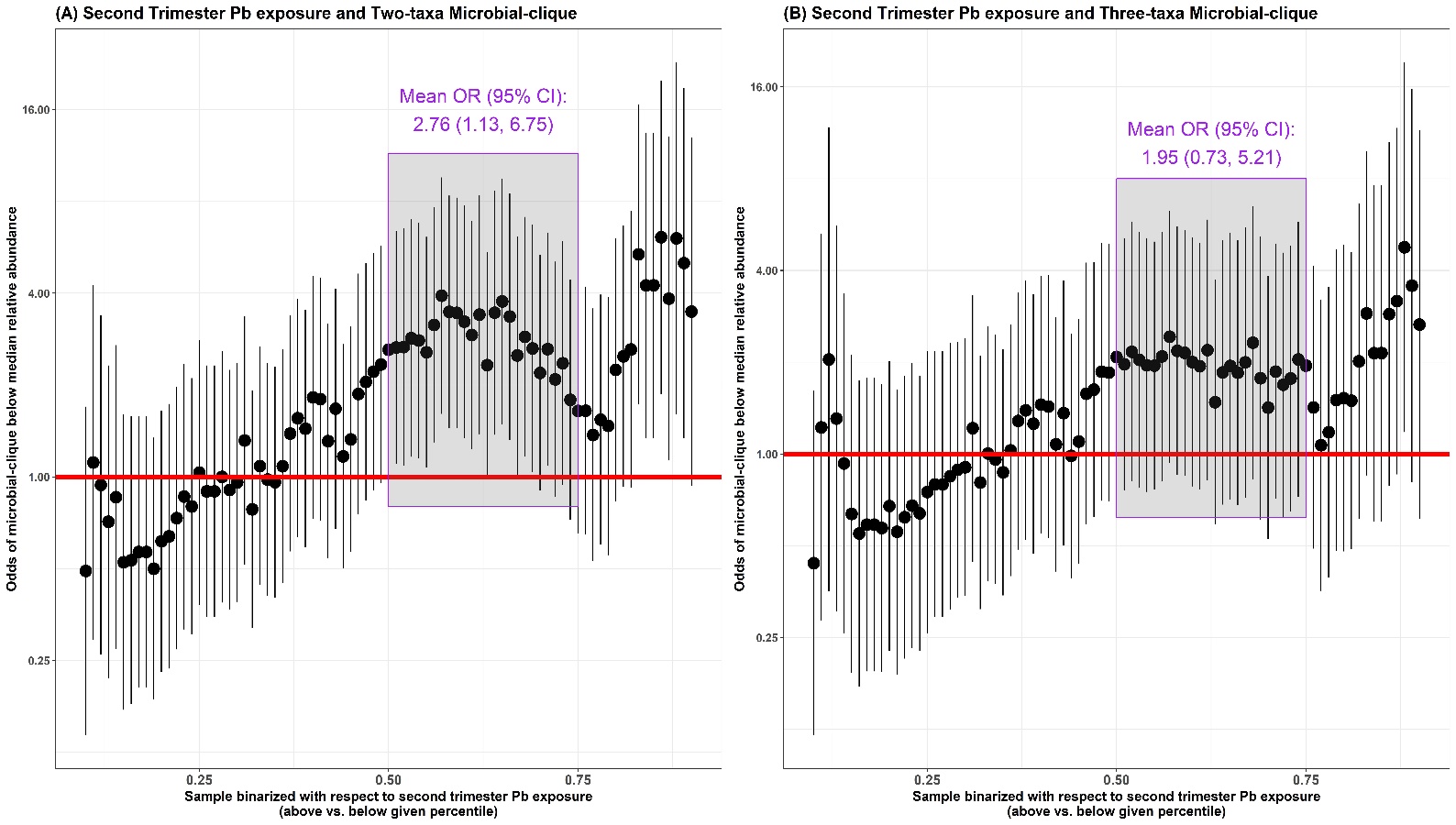

Odds of having below-median relative abundance of the (A) 2- and (B) 3-taxa cliques with respect to sample that was binarized using increasing quantiled 2T Pb concentration
